# Unraveling the genetic landscape of susceptibility to multiple primary cancers

**DOI:** 10.1101/2024.10.29.24316326

**Authors:** Pooja Middha, Linda Kachuri, Jovia L. Nierenberg, Rebecca E. Graff, Taylor B. Cavazos, Thomas J. Hoffmann, Jie Zhang, Stacey Alexeeff, Laurel Habel, Douglas A. Corley, Stephen Van Den Eeden, Lawrence H. Kushi, Elad Ziv, Lori C. Sakoda, John S. Witte

## Abstract

With advances in cancer screening and treatment, there is a growing population of cancer survivors who may develop subsequent primary cancers. While hereditary cancer syndromes account for only a portion of multiple cancer cases, we sought to explore the role of common genetic variation in susceptibility to multiple primary tumors. We conducted a cross-ancestry genome-wide association study (GWAS) and transcriptome-wide association study (TWAS) of 10,983 individuals with multiple primary cancers, 84,475 individuals with single cancer, and 420,944 cancer-free controls from two large-scale studies.

Our GWAS identified six lead variants across five genomic regions that were significantly associated (P<5×10^-8^) with the risk of developing multiple primary tumors (overall and invasive) relative to cancer-free controls (at 3q26, 8q24, 10q24, 11q13.3, and 17p13). We also found one variant significantly associated with multiple cancers when comparing to single cancer cases (at 22q13.1). Multi-tissue TWAS detected associations with genes involved in telomere maintenance in two of these regions (*ACTRT3* in 3q26 and *SLK* and *STN1* in 10q24) and the development of multiple cancers. Additionally, the TWAS also identified several novel genes associated with multiple cancers, including two immune-related genes, *IRF4* and *TNFRSF6B*. Telomere maintenance and immune dysregulation emerge as central, common pathways influencing susceptibility to multiple cancers. These findings underscore the importance of exploring shared mechanisms in carcinogenesis, offering insights for targeted prevention and intervention strategies.

## Introduction

Advances in cancer screening, diagnosis, and treatment have led to earlier detection of cancer and increased survival in cancer patients^1,2^. Given the substantial worldwide prevalence of cancer and the rising rates of survival, there has been a considerable increase in the number of cancer survivors who face an elevated risk of developing a second primary cancer during their lifetime^3–5^. Understanding risk factors for multiple primary cancers can have practical implications for managing patients with multiple cancers and may help prioritize screening strategies among cancer survivors. For patients with a history of cancer and prior anticancer treatments, distinguishing between a newly developed metastasis from the initial cancer and a second malignancy can be challenging and is under-studied^6^. Recognizing these scenarios and conducting appropriate investigations is essential for shaping subsequent therapeutic strategies. Furthermore, the presence of multiple primary cancers can impact a patient’s eligibility for enrollment in clinical research protocols, as individuals with a prior cancer history or concurrent secondary malignancies are typically excluded from clinical trials. Therefore, it is crucial to identify and understand risk factors associated with multiple primary cancers to manage and treat patients effectively.

Increased susceptibility to multiple cancers could be attributed to a variety of factors, such as lifestyle factors, environmental exposures, and genetic factors^3,4^. Furthermore, treatments such as radiation therapy or chemotherapy, which are often administered for the initial cancer diagnosis, introduce their own set of potential risks associated with the development of subsequent primary cancers^3,4^. Additionally, hereditary cancer syndromes, accounting for 1% to 2% of all people with cancer diagnoses, are associated with an elevated risk of multiple cancers^6^. Some studies have shown that individuals with multiple cancers have a higher frequency of deleterious variants in known cancer-risk genes^7–11^, and others have explored the cross-cancer effects of common germline genetic variants and reported associations with multiple variants in many specific genomic regions, such as 5p15.33, 6p21-22, 8q24, and 9q34^12–17^. Despite these known syndromes and genetic variants, much of the variability in the occurrence of multiple cancers remains unknown.

In this study, we examine the contribution of common germline genetic variants to the susceptibility of multiple primary cancers from a genome-wide perspective. Using data from two large studies, the UK Biobank (UKB)^18^ and Kaiser Permanente Northern California Genetic Epidemiology Research in Adult Health and Aging (GERA)^19^, we conducted a genome-wide association study (GWAS) and a transcriptome-wide association study (TWAS) to better understand the genetic basis and underlying biology associated with the development of multiple primary cancers.

## Materials and Methods

### Study populations and phenotyping

The study design and analyses are illustrated in Figure 1. The UKB is a population-based cohort comprising approximately 500,000 individuals aged 40-69 years recruited between 2006 and 2010 from various regions across the United Kingdom^18^. GERA is a prospective cohort consisting of nearly 102,979 adults who are members of the Kaiser Permanente Northern California (KPNC) health plan who participated in the Research Program on Genes, Environment, and Health (RPGEH). GERA included RPGEH participants who had provided samples for genotyping, had filled out a detailed health and lifestyle survey, and had detailed electronic health records available^19^.

**Figure 1:**
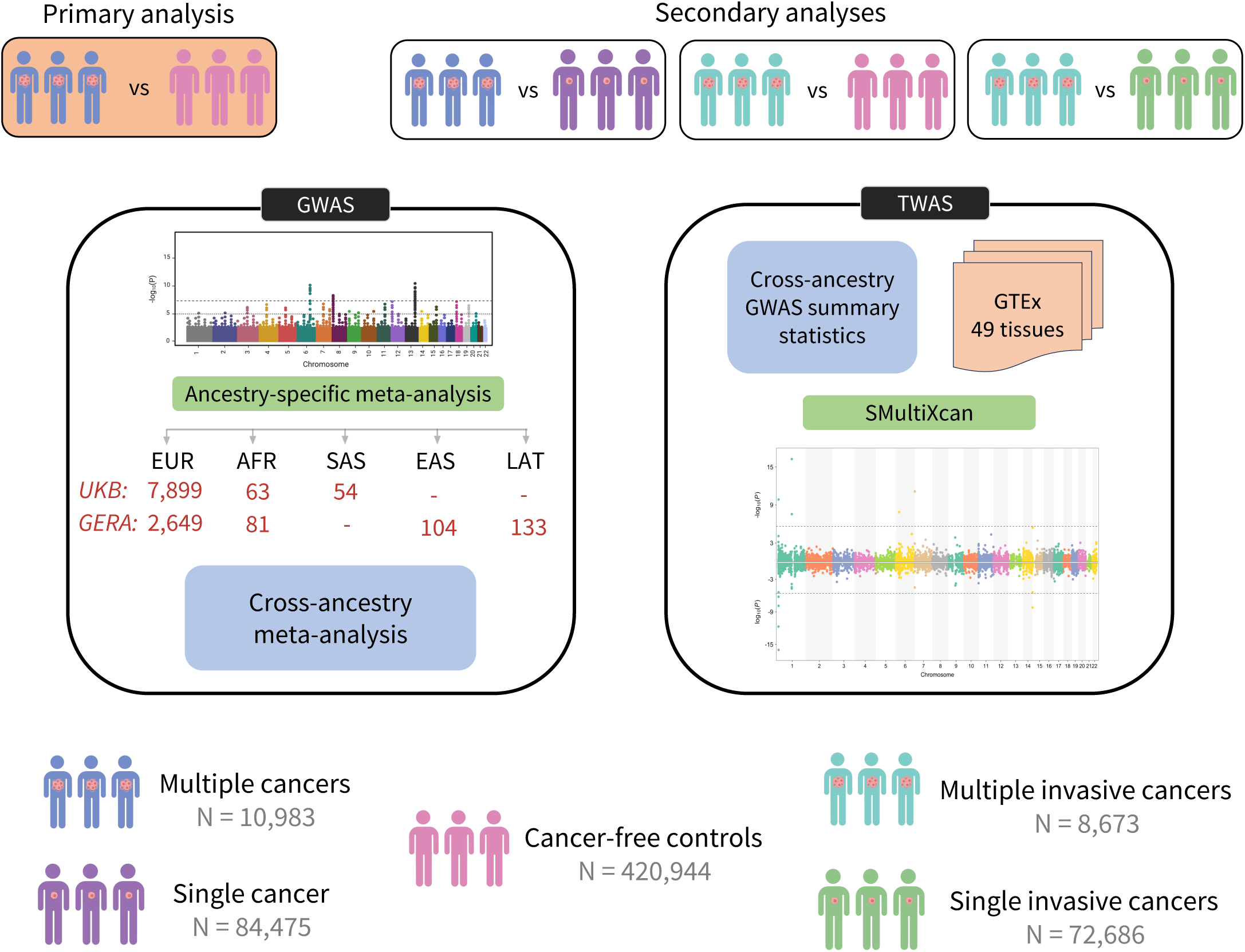
Schematic figure showing the study design and analytical pipeline of the study. Genome-wide association studies (GWAS) were performed in UK Biobank (UKB) and Kaiser Permanente Northern California Genetic Epidemiology Research in Adult Health and Aging (GERA) within each population grouping defined using a combination of self-reported race/ethnicity and clustering based on genetic ancestry principal components: European (EUR), African (AFR), South Asian (SAS), East Asian (EAS), and Latino (LAT). GWAS results from UKB and GERA were first combined within populations, using inverse-weighted fixed-effects meta-analysis, followed by a cross-ancestry meta-analysis. Next, using GTEx MASHR models from 49 tissues and cross-ancestry meta-analyzed GWAS summary results, we conducted multi-tissue transcriptome-wide association (TWAS) analyses. Primary analysis was performed for multiple cancer cases versus cancer-free controls (top left orange box). We conducted secondary analyses (top boxes with no background color): multiple cancer cases versus single cancer cases, multiple invasive cancer cases versus cancer-free controls, and multiple invasive cancer cases versus single invasive cancer cases. Sample size under the GWAS section corresponds to the number of multiple cancer cases by each population/ancestry group across UKB and GERA cohorts.

Cancer diagnoses and mortality data for UKB and GERA participants were obtained from national registries. Ascertainment of cancer diagnoses has been previously described^12,20^. Briefly, both studies included prevalent and/or incident diagnoses of invasive, borderline, and in situ primary tumors^20^. Non-melanoma skin cancer or metastatic cancer, as indicated by specific ICD codes, were not considered primary tumors. Most cancers were classified according to the SEER site recode paradigm^21^. We incorporated the morphological classifications outlined by the World Health Organization (WHO) for hematologic cancers, which categorized cancers into three major subtypes: lymphoid neoplasms, myeloid neoplasms, and NK- and T-cell neoplasms^22^.

Multiple cancer cases were defined as individuals with ICD-9 or ICD-10 codes indicating primary tumors at two or more distinct organ sites. Single cancer cases were defined as individuals with ICD-9 or ICD-10 codes indicating a primary tumor at only one organ site. Multiple and single invasive cancer cases were limited to participants with primary invasive cancers and excluded those with borderline and in-situ malignancies. In the UKB, controls were all individuals without a cancer diagnosis at the last follow-up. In the GERA cohort, cancer-free controls at the last follow-up were matched 1:1 to case individuals based on age at specimen collection, sex, genotyping array, and reagent kit.

All participants in the study provided informed consent. The UKB obtained ethical approval from the Research Ethics Committee, adhering to the UKB Ethics and Governance Framework. Approval for the original GERA study was granted by the institutional review board of Kaiser Permanente Northern California and the Human Research Protection Program (Committee on Human Research) at the University of California, San Francisco.

### Genotyping and quality control

Genotyping and imputation processes for the UKB cohort have been previously described^12,20^. In brief, participants were genotyped on the UKB Affymetrix Axiom array (89%) or the UK BiLEVE array (11%)^18^. Imputation was performed using the Haplotype Reference Consortium (HRC), along with merged reference panels from UK10K and 1000 Genomes phase 3^18^. Genetic ancestry principal components (PCs) were computed using fastPCA, based on a set of 407,219 unrelated samples and 147,604 genetic markers^18^. Population groupings for GWAS in UKB were determined based on self-reported race/ethnicity and genetic ancestry PCs. Individuals for whom either of the first two ancestry PCs fell outside of five standard deviations from the population mean within each self-reported ethnicity group were excluded from the analyses^12^. Using a robust method KING, we excluded samples with discordant self-reported and genetic sex and one individual from each pair of first-degree relatives^12,23^. Using a subset of genotyped autosomal variants with MAF□≥□1% and call rate□≥□97%, we excluded samples with heterozygosity >5□standard deviations from the mean^18^.

For the GERA cohort, genotyping was conducted using one of four Affymetrix Axiom arrays optimized for African, East Asian, European, and Latino racial/ethnic groups^24,25^, and has been previously described in detail^24–26^. Briefly, population groupings were determined based on self-reported race/ethnicity and PCs. Imputation was conducted using SHAPE-ITv2.565^27^ for pre-phasing genotypes, followed by IMPUTE2 v2.3.1^28^ using the 1000 Genomes Project Phase I as a cosmopolitan reference panel^28^. Ancestry principal components (PCs) were estimated with Eigenstrat v4.2, as has been described^29,30^.

Analyses in both UKB and GERA were restricted to population groups with at least 50 multiple cancer individuals. Additional quality control (QC) measures were applied at variant level. Specifically, variant-level QC filters included imputation quality (INFO□<□0.30), minor allele frequency (MAF<0.01 in European population group and MAF<0.05 in African, East Asian, South Asian, and Latinx population groups).

### Statistical Analysis

The study design and analytical strategy of our study is illustrated in Figure 1. The primary GWAS analysis compared participants with multiple cancer diagnoses to those without cancer in UKB and GERA within each population group defined using a combination of self-reported race/ethnicity and clustering based on genetic ancestry PCs. In the UKB analysis, all models were adjusted for age, sex, the first ten global PCs, and the genotyping array. In the GERA study, all models were adjusted for age, sex, and genotyping array, and 10 PCs within the European group or 6 PCs within each of the other population groups. GWAS results from UKB and GERA were first combined within each population group using inverse-variance-weighted fixed-effects meta-analysis using METAL, followed by cross-ancestry meta-analysis^31^. Cross-ancestry meta-analysis results were clumped around index variants with the lowest genome-wide significant (P<5×10^−8^) meta-analysis p-value to identify independent association signals. Clumping was performed and variants with linkage disequilibrium (LD) r^2^□>□0.01 within a ±10- Mb window of the lead variant were assigned to that lead variant.

Next, we performed a multi-tissue transcriptome-wide association study (TWAS) using the cross-ancestry meta-analyzed summary statistics and S-MultiXcan framework to estimate the association between imputed gene expression and multiple cancers^32,33^. We used the multivariate adaptive shrinkage prediction models from 49 tissues in GTEx version 8^34^. An advantage of this approach is that it leverages the correlation in cis-eQTL effects across tissues. Genes with P<2.3×10^-6^ (0.05/22,244 genes Bonferroni correction) were considered statistically significant.

In addition to the primary analyses comparing cases with multiple cancers versus cancer-free controls, secondary GWAS and TWAS analyses were performed comparing: 1) a more restrictive case definition that restricted to multiple invasive neoplasms (excluding in-situ tumors) versus cancer-free controls; 2) cases with multiple cancers versus cases with a single cancer (case-case); and 3) cases with multiple invasive cancers versus cases with a single invasive cancer (case-case). Analyses were conducted using Plink2 and R v4.2.2 (R Foundation for Statistical Computing).

## Results

Characteristics of the study populations are shown in Table 1. In total, the studies included 10,983 individuals with multiple primary cancers (of which 8,673 had invasive cancers), 84,475 individuals with single cancers (72,686 with single invasive cancers), and 420,944 cancer-free controls. Among all multiple cancer cases, there were a total of 387 distinct cancer pairs. Of these, 47 cancer pairs had at least 50 individuals with the pair of cancers (Figure 2). The top pairs represented were cervix and breast (N=444), followed by prostate and melanoma (N=303), and bladder and prostate (N=302). Stratified by the study cohort, the top pairs in UKB were cervix and breast (N=387), bladder and prostate (N=249), and breast and colorectal (N=218) (Supplementary Figure 1). The top pairs in GERA were prostate and melanoma, with prostate cancer being the first cancer (N=177), melanoma and prostate where melanoma was the first cancer (N=125), and prostate and bladder (N=118) (Supplementary Figure 1).

**Figure 2:**
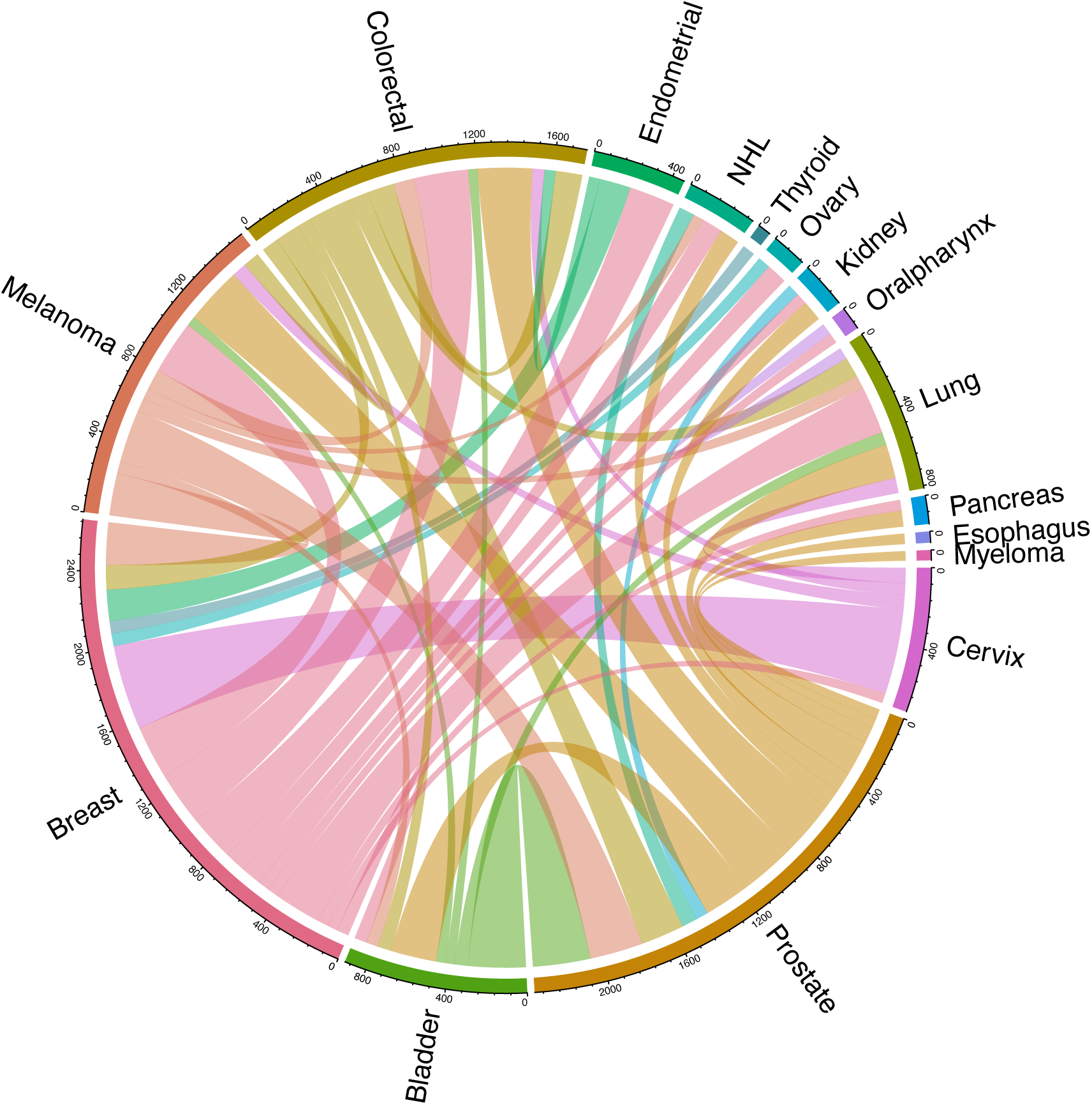
Circos plot showing the pairs of cancer diagnoses with at least 50 individuals in the UK Biobank and GERA studies combined. Each connection reflects the number of people with both linked primary cancers, where the color of the line shows the first cancer site diagnosed.

**Table 1:** Characteristics of the study population by cohort.

| Characteristics | UK Biobank |  |  | GERA |  |  |
| --- | --- | --- | --- | --- | --- | --- |
|  | Multiple Cancers<br>(N=8,016) | Single Cancer<br>(N=66,410) | Controls<br>(N=355,100) | Multiple Cancers<br>(N=2,967) | Single Cancer<br>(N=18,065) | Controls<br>(N=65,844) |
| **Population/**<br>**Ancestry**<br>**groups<sup>#</sup>** |  |  |  |  |  |  |
| African | 63<br>(1) | 784<br>(1) | 6,394<br>(2) | 81<br>(3) | 519<br>(3) | 2,327<br>(4) |
| East Asian | - | - | - | 104<br>(4) | 993<br>(5) | 5,737<br>(9) |
| European | 7,899<br>(98) | 64,885<br>(98) | 340,519<br>(96) | 2,649<br>(89) | 15,425<br>(85) | 51,456<br>(78) |
| Latino | - | - | - | 133<br>(4) | 1,128<br>(6) | 6,324<br>(10) |
| South Asian | 54<br>(1) | 741<br>(1) | 8,187<br>(2) | - | - | - |
| **Age at**<br>**Assessment<sup>\*</sup>** | 62<br>(6) | 60<br>(7) | 56<br>(8) | 72<br>(10) | 68<br>(11) | 61<br>(14) |
| **Female<sup>#</sup>** | 4,318<br>(54) | 37,805<br>(57) | 188,475<br>(53) | 1,439<br>(49) | 9,987<br>(55) | 38,917<br>(59) |
| **Age at First**<br>**Cancer<sup>\*</sup>** | 56<br>(13) | 59<br>(12) | - | 64<br>(12) | 65<br>(13) | - |
| **Age at Second**<br>**Cancer<sup>\*</sup>** | 65<br>(10) | - | - | 73<br>(11) | - | - |

<sup>#</sup> : Number (Percentage)
<sup>\*</sup> : Mean (Standard deviation)
Age at assessment was defined as age at diagnosis for cases and age at enrollment for controls

### Genetic variants associated with multiple cancers

We conducted a cross-ancestry genome-wide association study (GWAS) and meta-analysis of 10,983 individuals with multiple primary tumors and 420,944 cancer-free controls. We refer to this as our primary analysis. Additionally, we conducted a sensitivity analysis restricting to 8,673 individuals with multiple invasive tumors only, excluding in-situ diagnoses. To isolate genetic susceptibility specifically for multiple primary cancers, we carried out additional analyses comparing individuals with multiple cancers to those with any single primary cancer (N=84,475 cancer controls). Parallel analyses were conducted restricting the comparison group to individuals with single primary invasive cancers (N=72,686 single invasive cancer controls) (Figure 1).

Our primary GWAS identified four independent genome-wide significant variants across four genomic regions associated with the risk of developing multiple cancers (Table 2 and Figure 3). Of the four lead genomic regions in Table 2, three were genome-wide significant in European ancestry individuals: rs2293607, rs201581170, and rs612611, one was genome-wide significant in African ancestry individuals: rs72725854. None were significant in the other population groups (Table 2). In the analyses restricted to 8,673 participants with multiple invasive cancers and 420,944 cancer-free controls, we identified three SNPs (3 European) across three loci (rs283732, rs9419958, andrs35850753) associated with multiple invasive cancers at P<5×10^-8^ (Supplementary Table 2, Supplementary Table 3 and Supplementary Figure 2). To further characterize genetic susceptibility to multiple primary cancers and distinguish these signals from general cancer risk loci, we conducted case-case comparisons. None of the variants identified in our case-control analyses reached genome-wide significance when comparing multiple cancers to single-cancer controls. However, we found one genome-wide significant genetic variant in the 22q13.1 region (rs192493667, OR=1.39, P=3.6×10^-8^) in our multiple primary cancers and single cancer control analysis. The association for this variant was driven by the European population group. Parallel case-case GWAS that was restricted to invasive tumors did not identify any genome-wide significant associations.

**Figure 3:**
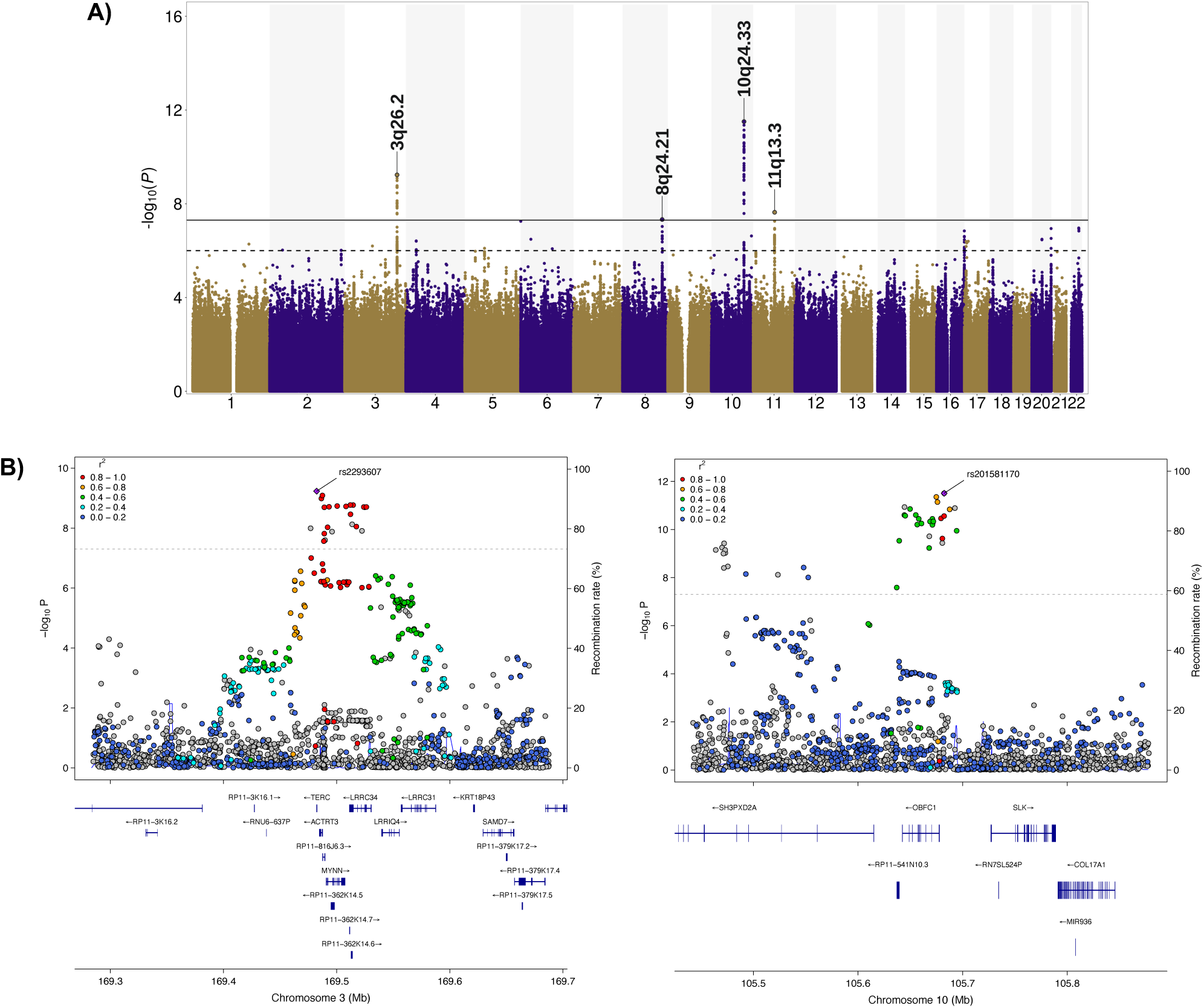
Manhattan plot (A) highlighting the lead chromosomal regions (P<5×10^-8^) from the genome-wide association study of multiple cancers (10,983 multiple cancers, 420,944 cancer-free controls). The solid black line signifies the genome-wide significance threshold (P<5×10^-8^), whereas the dashed black represents a suggestive significance threshold of P<1×10^-6^. (B) Locus zoom plots map the genomic location of significant SNPs on chr 3 and 10 to the *ACTRT3 and OBFC1* loci, respectively, with the lead SNP annotated.

**Table 2:** Lead variant from 4 genomic regions (P<5×10^-8^) with meta-analyzed and ancestry-specific estimate from the genome-wide association study of multiple cancers and cancer-free controls.

| Region | Chr | Position | SNP | E/O | EAF<sub>Meta</sub> | OR<sub>Meta</sub> | P<sub>Meta</sub> | OR<sub>EUR</sub> | P<sub>EUR</sub> | OR<sub>AFR</sub> | P<sub>AFR</sub> | OR<sub>SAS</sub> | P<sub>SAS</sub> | OR<sub>EAS</sub> | P<sub>EAS</sub> | OR<sub>LAT</sub> | P<sub>LAT</sub> | P<sub>HET</sub> |
| --- | --- | --- | --- | --- | --- | --- | --- | --- | --- | --- | --- | --- | --- | --- | --- | --- | --- | --- |
| 3q26.2 | 3 | 169482335 | rs2293607 | T/C | 0.75 | 1.11 | $5.8 \times 10^{-10}$ | 1.10 | $4.5 \times 10^{-9}$ | 1.07 | 0.78 | 1.48 | 0.08 | 1.47 | 0.009 | 0.99 | 0.99 | 0.2 |
| 8q24.21 | 8 | 128074815 | rs72725854 | A/T | 0.92 | 0.28 | $4.7 \times 10^{-8}$ | - | - | 0.28 | $4.7 \times 10^{-8}$ | - | - | - | - | - | - | 1 |
| 10q24.33 | 10 | 105682344 | rs201581170 | A/T | 0.16 | 1.16 | $3.1 \times 10^{-12}$ | 1.15 | $2.2 \times 10^{-11}$ | 1.30 | 0.2 | 1.65 | 0.02 | - | - | - | - | 0.22 |
| 11q13.3 | 11 | 69307463 | rs612611 | A/G | 0.85 | 0.90 | $2.3 \times 10^{-8}$ | 0.90 | $4 \times 10^{-8}$ | 0.92 | 0.64 | 0.85 | 0.51 | - | - | 0.89 | 0.53 | 0.99 |

Chr: Chromosome, SNP: Single nucleotide polymorphism, E/O: Effect allele/Other allele, EAF: Effect allele frequency, OR: Odds ratio, Meta: Meta-analysis, EUR: European, AFR: African, SAS: South Asian, EAS: East Asian, LAT: Latino, P<sub>HET</sub>: P-value for heterogeneity

The lead variant in the 3q26.2 region, rs2293607 (*TERC*), was associated with an increased risk of multiple cancers (OR=1.11, P=5.8×10^-10^), and the magnitude of the association was comparable when restricting to invasive tumors (OR=1.10, P=1.5×10^-7^), but decreased considerably when cancer-free controls were replaced with single cancer controls (OR=1.06, P=0.001). Next, we identified two susceptibility signals in the 10q24.33 region. The first lead variant, rs201581170, was associated with an increased risk of multiple cancers (OR=1.16, P=3.1×10^-12^; invasive: OR=1.17, P=2.4×10^-10^) and remained directionally consistent in case-case analyses (OR=1.10, P=4.2×10^-5^; invasive: OR=1.10, P=3×10^-4^) (Supplementary Table 1). The second risk variant, rs9419958 (*STN1*), was detected when comparing multiple invasive cancers to cancer-free controls (OR=1.16, P=1.7×10^-11^). This SNP is in high LD with rs201581170 (r^2^=0.83) and also had an association with multiple cancers (OR=1.14, P=7.2×10^-12^). However, its effect on risk was weakened when cancer-free controls were replaced with individuals diagnosed with a single invasive cancer (OR=1.09, P=1.4×10^-4^) (Supplementary Table 3).

We found two SNPs in the 8q24.21 region. The first, rs72725854 showed a significant association with a decreased risk of multiple cancers (rs72725854: OR=0.28, P=4.7×10^-8^). The association remained consistent in analyses restricted to multiple invasive cancers (OR=0.26, P=1.3×10^-7^) and when compared to single cancer controls (OR=0.33, P=6.4×10^-5^). Another SNP, rs283732, demonstrated a significant association with a decreased risk of multiple invasive cancers (OR=0.90, P=1.1×10^-8^). This SNP is not in LD with rs72725854 (r^2^=0.003). We also found an inverse association between rs612611 (11q13.3) and the risk of multiple cancers (OR=0.90, P=2.3×10^-8^). The association for both rs283732 and rs612611 was stable but not significant across all analyses (Supplementary Table 1 and Supplementary Table 3).

Lastly, we found a risk signal for multiple invasive cancers associated with rs35850753, an intronic variant in *TP53* (OR=1.35, P=1.2×10^-8^), which exhibited a slightly weaker association when in-situ tumors were also included (OR=1.27, P=4.7×10^-7^). The magnitude of effect for rs35850753 was slightly attenuated in case-case analyses (OR=1.16, P=0.003; invasive: OR=1.24, P=8.1×10^-5^). Case-case analysis with single cancer as controls reported a genome-wide significant association between rs192493667 in the 22q13.1 region and multiple cancers (OR=1.39, P=3.6×10^-8^). This increased risk between rs192493667, and multiple cancers was consistent across all analyses.

### TWAS identifies genes involved in telomere maintenance and immune dysregulation

We undertook a multi-tissue TWAS to assess the associations between genetically predicted gene expression and the risk of developing multiple cancers, following the same case-control and case-case comparison framework as the GWAS. Of the 22,244 genes tested, statistically significant (Bonferroni correction: P<2.3×10^-6^) associations were detected for 9 genes across 8 genomic loci (Table 3 and Figure 4). The TWAS recapitulated some of the loci identified in the GWAS, such as *ACTRT3* in 3q26 and *SLK* and *STN1* in 10q24 (Figure 1b and Figure 4a). Genetically predicted expression of *LMAN2L* (2q11, P=5.3×10^-7^), *ACTRT3* (3q26, P=1.8×10^-8^), *IRF4* (6p25, P=1.8×10^-6^), *FAM208B* (10p15, P=1.9×10^-7^), *SLK* (10q24, P=4.6×10^-8^), *STN1* (10q24, P=1×10^-6^), *SPIRE2* (16q24, P=6.5×10^-7^), and *TNFRSF6B* (20q13, P=6×10^-10^) were associated with risk of developing multiple cancers, and *ARFGAP3* (22q13, P=1.9×10^-6^) was associated with multiple invasive cancers. Supplementary Table 4 reports the associations of these genes with multiple cancers in each tissue where the models were available. The case-case TWAS found four genes associated with multiple cancers: *PSMA2*, *PLOD1*, *CDH5*, and *PLEKHM1* (Supplementary Table 5 and Supplementary Figure 3).

**Figure 4:**
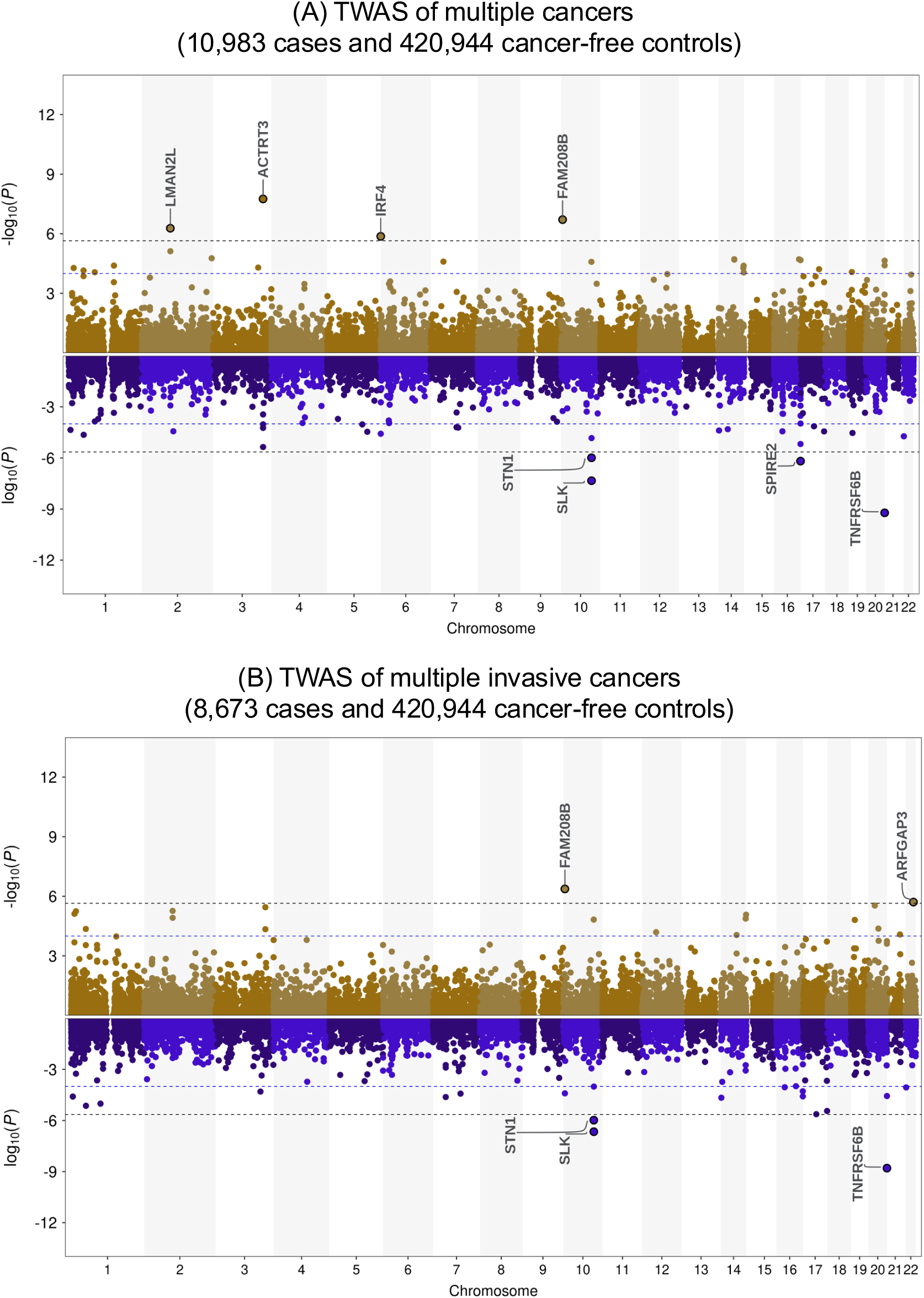
Multi-tissue transcriptome-wide association study (TWAS) results for (A) multiple cancers and cancer-free controls and (B) multiple invasive cancers and cancer-free controls. The top panel of the Miami plots depicts the associations for genes with mean Z scores > 0, and the bottom panel shows genes with mean Z scores ≤ 0. The threshold for statistical significance was determined based on the Bonferroni correction for 22,244 genes tested (P< 2.3×10^-6^, dashed black line), while the suggestive significance threshold was set at P<1×10^-4^ (dashed blue line).

**Table 3:** Statistically significant genes (P<0.05/22,244), highlighted in bold) identified from the multi-tissue transcriptome-wide association study (TWAS) comparing multiple cancer cases to cancer-free controls, and multiple invasive cancer cases to cancer-free controls. The table also presents the associations of these significant genes in additional related analyses

| Region | Gene name | Multiple cancer and cancer-free controls |  | Multiple invasive cancers and cancer-free controls |  | Multiple cancers and single cancer as controls |  | Multiple invasive cancers and single invasive cancer as controls |  |
| --- | --- | --- | --- | --- | --- | --- | --- | --- | --- |
|  |  | Z (mean) | P value | Z (mean) | P value | Z (mean) | P value | Z (mean) | P value |
| 2q11.2 | *LMAN2L* | 3.58 | **5.3×10<sup>-7</sup>** | 3.27 | 5.4×10<sup>-6</sup> | 3.16 | 2.1×10<sup>-5</sup> | 2.95 | 7×10<sup>-5</sup> |
| 3q26.2 | *ACTRT3* | 0.93 | **1.8×10<sup>-8</sup>** | 0.82 | 3.6×10<sup>-6</sup> | 0.13 | 5.4×10<sup>-3</sup> | 0.11 | 2.1×10<sup>-2</sup> |
| 6p25.3 | *IRF4* | 3.77 | **1.5×10<sup>-6</sup>** | 2.95 | 2.8×10<sup>-4</sup> | 2.05 | 2.3×10<sup>-2</sup> | 1.57 | 0.14 |
| 10p15.1 | *FAM208B* | 1.06 | **1.9×10<sup>-7</sup>** | 1.03 | **4.3×10<sup>-7</sup>** | 0.62 | 1×10<sup>-3</sup> | 0.61 | 1.2×10<sup>-3</sup> |
| 10q24.33 | *SLK* | -0.66 | **4.6×10<sup>-8</sup>** | -0.32 | **2.2×10<sup>-7</sup>** | -0.53 | 3.1×10<sup>-2</sup> | -0.18 | 3.1×10<sup>-2</sup> |
| 10q24.33 | *STN1* | -1.17 | **1×10<sup>-6</sup>** | -1.48 | **1×10<sup>-6</sup>** | 0.06 | 4.5×10<sup>-4</sup> | -0.42 | 1.5×10<sup>-3</sup> |
| 16q24.3 | *SPIRE2* | -1.56 | **6.5×10<sup>-7</sup>** | -1.85 | 2.6×10<sup>-5</sup> | -1.19 | 8.2×10<sup>-3</sup> | -1.42 | 2.3×10<sup>-2</sup> |
| 20q13.33 | *TNFRSF6B* | -1.74 | **6×10<sup>-10</sup>** | -2.01 | **1.6×10<sup>-9</sup>** | -1.27 | 1.9×10<sup>-3</sup> | -1.62 | 2.3×10<sup>-3</sup> |
| 22q13.2 | *ARFGAP3* | 0.32 | 1.1×10<sup>-4</sup> | 0.21 | **1.9×10<sup>-6</sup>** | 0.24 | 2.3×10<sup>-3</sup> | 0.06 | 6.8×10<sup>-5</sup> |

The signal in the 3q26 region involved a cluster of genes: *ACTRT3*/*APRM1*, *MYNN*, *LRRC34*, and *LRRIQ4*, but only *ACTRT3* was associated at a Bonferroni P<2.3×10^-6^. Elevated expression of *ACTRT3* was associated with the development of multiple cancers across 15 tissues, with three tissues (cerebellar hemisphere, cerebellum, tibial nerve) showing an inverse association. However, *ACTRT3* expression was not significantly associated with risk of multiple cancers (overall and invasive) in either of the case-case analyses.

For *IRF4*, predicted expression models were found in 33 different tissues, and they showed significant associations with increased cancer susceptibility in 20 of those tissues. Interestingly, all 20 tissues exhibited a positive association, except for sun-exposed lower leg skin. The expression of *IRF4* showed a similar trend toward the risk of developing multiple cancers (overall and invasive) across all secondary analyses; however, none were significant after accounting for multiple testing correction. *TNFRSF6B* expression showed a significant inverse association with susceptibility to multiple cancers across five tissues, including EBV-transformed lymphocyte cells and prostate. In other tissues, the association of increased *TNFRSF6B* expression with multiple cancers was heterogeneous and not significant. In our multiple invasive cancer and cancer-free controls TWAS analysis, we identified a significant association between increased expression of *TNFRSF6B* and multiple invasive cancers (P=1.6×10^-9^). None of the associations were significant when restricted to case-case analyses.

Two genes in the 10q24 region showed association with susceptibility to multiple cancers. We identified an inverse association between elevated expression of *SLK* and multiple cancers across 31 tissues out of 44 tissues with a model available for this gene. The remaining 13 tissues, including ovary, prostate, and whole blood, showed a positive association between *SLK* expression and multiple cancers. Results for *STN1* were mostly consistent across the tissues with decreased expression of *STN1* associated with susceptibility to multiple cancers except in arterial tissues, neurological tissues, prostate, ovary and kidney cortex. This inverse association between increased *STN1* expression and multiple cancers was significant in cerebellar hemispheres, esophagus muscularis, and thyroid. The association with the expression of *SLK* (P=2.2×10^-7^) and *STN1* (P=1×10^-6^) was also significantly associated with multiple invasive cancers.. However, the associations between multiple cancers (overall and invasive) and expression of *SLK* and *STN1* were not significant in either of the case-case analyses.

## Discussion

Advances in cancer screening, prevention, and treatment have improved survival rates, leading to an increasing number of cancer survivors who may face an elevated risk of developing a second primary cancer^3,6^. Understanding the potential causes and risk factors of second primary cancers is thus crucial to managing and treating patients effectively. In this study, we sought to investigate the role of germline genetics on the risk of multiple primary cancers through GWAS and TWAS. Our GWAS findings revealed associations of common germline variants in several genomic regions, including 3q26, 8q24, 10q24, and 17p13, with multiple cancers. Previous work has shown that these genomic regions and genetic variants are well-established as cancer susceptibility loci,^35–42^ and that some of the variants have previously reported to have pleiotropic effects^12–17,20^.

Our multi-tissue TWAS recapitulated some of these regions: *ACTRT3* (3q26), and *SLK*, *STN1* (10q24). We found an immune-related gene, *TNFRSF6B*, associated with the risk of developing multiple cancers. Additionally, *IRF4* has previously been implicated in single individual cancers^43–47^. However, our findings suggest that *IRF4* may have pleiotropic effects, influencing the susceptibility to multiple cancers. Overall, the findings of this study reiterate the complex role of telomere maintenance and immunological pathways in the development of multiple primary tumors. Understanding these genetic contributions could improve risk stratification and inform screening and prevention strategies for individuals at risk of developing second primary cancers.

The GWAS findings of associated SNP rs2293607 in 3q26.2 alongside TWAS associations suggest that this region may influence cancer risk through transcriptional regulation, which may lead to pleiotropic effects across different cancer types. Interestingly, rs2293607, has been previously implicated in diverse phenotypes, including longer leukocyte telomere length^48,49^ and cancer pleiotropy^12,13,50^. Further exploration of this region revealed that SNPs in strong LD with rs2293607 (r² ≥ 0.90) also demonstrated associations with longer telomere length^49,51–54^ and were implicated in susceptibility to multiple cancers^19,54,55^, including colorectal cancer^42,53,56^, bladder cancer^57–59^, multiple myeloma^60,61^, lung adenocarcinoma^62^, and thyroid cancer^36,63^. Associated variants within the 3q26 region map to the *TERC* gene cluster, which includes *ACTRT3*/*APRM1*, *MYNN*, *LRRC34*, and *LRRIQ4*^64^. The genetic variants within the *TERC* gene cluster have been previously implicated in promoting longer telomere length through telomerase over-expression^49,51–54,65–67^. Genetic predisposition to telomere maintenance is believed to promote cancer development by fostering continuous cell proliferation and the accumulation of mutations. In addition to increasing risk of specific cancers, such as lung^68,69^, melanoma^68,70^, and glioma^71^, our results indicate that genetic mechanisms resulting in longer telomere length may also influence predisposition to multiple primary tumors. Our TWAS detected a Bonferroni-significant positive association between the genetically predicted expression of *ACTRT3* within the *TERC* gene cluster and susceptibility to multiple cancers. While the associations of *MYNN*, *LRRC34*, and *LRRIQ4* did not meet our significance threshold, we did observe a positive association between *LRRC34* and the diagnosis of multiple cancers, along with an inverse association between *LRRIQ4* and *MYNN* and multiple cancer diagnoses. These associations between each gene and multiple cancer diagnoses across tissues, which is in sync with the fact that telomere length varies across tissue types^72^. Understanding these genetic underpinnings could contribute valuable insights into the molecular pathways involved in cancer development, offering new avenues for targeted interventions and therapeutic strategies.

The 10q24.33 region has shown a notable effect in the context of cancer susceptibility^12,20^, with multiple SNPs within the locus being associated with an increased risk of both multiple cancers and multiple invasive cancers. The SNP rs201581170 demonstrated a consistent direction of effect (although not reaching genome-wide significance in case-case analyses), suggesting a broader role of this SNP in the development of multiple cancer types, indicative of pleiotropic behavior across malignancies. Furthermore, rs9419958, which is in high LD with rs201581170, was associated with multiple invasive cancers. We believe that these two SNPs are capturing the same signal in relation to multiple cancer susceptibility. These findings are further supported by the TWAS analyses, with two genes, *SLK* and *STN1*, showing significant associations with multiple cancers across several tissues. *STN1 gene* also known as *OBFC1* gene encodes a component of the CST (CTC1-STN1-TEN1) complex^73^. ^48^. This complex is essential for the maintenance of telomere ends, ensuring genomic stability, which is a critical factor in preventing replicative senescence and the onset of malignancies^73^. The association between telomere length and cancer susceptibility, mediated by variants in *OBFC1* is well known40,49, 65, 70, 71. While telomere length may confer risk for cancer in some tissues, the impact may vary across different tissue types due to variations in cellular turnover rates, telomerase activity, and other tissue-specific factors72^74^., We also found a SNP rs35850753 associated with the increased risk of multiple invasive cancers. This intronic SNP is located in the 5’-UTR of *TP53*, which is known to increase the risk of many cancers, including breast, lung, leukemia, and neuroblastoma^75–78^.

Our TWAS also identified a positive association between *IRF4* expression and multiple primary cancers. This association was consistent across most tissues. This gene has been previously associated with skin cancer^43,79,80^, lung cancer^47^, and hematological malignancies^44,80,81^. *IRF4* is a member of the interferon regulatory factor (IRF) family of transcription factors, predominantly expressed in immune cells, where it transduces signals from various receptors to activate or repress gene expression^82,83^. *IRF4* plays a crucial role in regulating various stages of lymphoid, myeloid, and dendritic cell differentiation^84^. Given the ubiquitous involvement of inflammation in carcinogenesis, *IRF4* is an important cancer susceptibility gene because of its role as a regulator of immune response. Experimental studies of non-small cell lung cancer cell lines have demonstrated that overexpression of *IRF4* exhibits tumor promoter activity, partially attributable to the activation of the Notch-Akt signaling pathway^45^. Furthermore, the implications of *IRF4* extend beyond experimental settings, as its expression has been identified as a prognostic factor in various malignancies, including hematological malignancies^46^, lung cancer^85^, and breast cancer^86^.

Our cross-tissue TWAS analysis showed an inverse association between *TNFRSF6B* in 20q13.3 and the risk of multiple cancers. This association was heterogeneous across tissues with most tissues showing an inverse association. However, tissues from colon, breast, spleen, uterus, liver, and ovary showed a positive association. This correlates with previously published studies where elevated expression of the gene has been implicated in diverse tumors, such as colorectal^87–90^, pancreatic^91–94^, ovarian^95–97^, and liver cancers^87,98,99^. Overexpression of *TNFRSF6B* promotes the immune evasion of tumor cells and inhibits apoptosis^100^. Acting as a decoy, *TNFRSF6B* competitively binds with FasL ligands, expressed on the T cells, impeding their ability to induce apoptosis and eliminate tumor cells^100^. This contributes to the development of tumors. Additionally, *TNFRSF6B* influences the differentiation of immune cells such as T lymphocytes, macrophages, and dendritic cells, thereby causing immune dysregulation and promoting tumor angiogenesis^100–102^. Thus, the dysregulation of the immune system via *TNFRSF6B* across various tissues emphasizes its potential involvement in tumorigenesis and indicates its possible role in the development of multiple cancers.

We also observed a significant association between the expression of the *PSMA2* gene in the 7p14 region and the risk of developing multiple cancers in our case-case analyses. Although *PSMA2* was not significantly associated with multiple cancers when comparing to cancer-free controls, the finding from the case-case analysis suggests that this gene may have a role in cancer development. *PSMA2* is an essential component of the 20S proteasome and impacts the proteasomal activity, which is crucial to tumor growth and immune evasion^103–107^. The role of elevated *PSMA2* expression has been implicated in cancer progression by enhancing cellular proliferation, migration, and invasion across several tumor types, including colorectal, breast, and lung^103,105,106^.

This is the first study to investigate the role of germline genetics on the susceptibility to multiple cancers from a genome-wide perspective. The study has several strengths that contribute to the robustness of our findings. First, the incorporation of two distinct comparison groups, namely cancer-free and single-cancer individuals, allows us to discern signals specific to pleiotropic effects versus signals for the independent development of multiple cancers. Another strength of this study is that UKB and GERA are both linked to cancer registries and, therefore, ensure high-quality ascertainment of incident cancer diagnoses. Further, the inclusion of participants from diverse ancestry groups in the two large cohorts bolsters the generalizability of our results. However, our study also has certain limitations. First, combining multiple primary cancers into a single phenotype does not consider the timing and sequence of specific cancer diagnoses and, therefore poses a challenge for inferring the mechanisms underlying the observed associations. We were also unable to account for screening behaviors (which may influence cancer prevention, detection, and stage at diagnosis) and for the type of cancer treatment administered after initial cancer diagnosis, which may be an independent risk factor for developing a second primary tumor. Finally, our power to detect loci with small effect sizes in non-European populations may have been limited due to small sample sizes in these groups. Future research should prioritize larger studies in diverse populations, ensuring a more comprehensive understanding of the genetic underpinnings of multiple cancers.

Overall, our study investigates the genetic etiology of multiple primary cancers, unveiling the presence of pleiotropic signals that transcend traditional single-cancer associations. The convergence of genetic variants within known cancer pleiotropic regions suggests a shared molecular basis underlying diverse cancer types, including telomere maintenance. Additionally, we have identified two immune-related genes, *IRF4*, and *TNFRSF6B,* associated with the diagnosis of multiple primary cancers, highlighting the potential contribution of immune dysregulation to this genetic landscape, and emphasizing common pathways influencing the susceptibility to multiple cancers. These findings emphasize the need for research into the molecular mechanisms underlying development of multiple cancers for informing potential interventions and targeted therapies.

## Data Availability

The UK Biobank is an open-access data resource, and the data is available from the UK Biobank access portal at https://www.ukbiobank.ac.uk. This research was conducted with approved access to UK Biobank data under application number 14105. The Kaiser Permanente Research Bank data are available via dbGaP (phs002809.v1.p1). Results generated from this work are included in the published article or Supplementary Materials. All additional data corresponding to the findings of this study are available within the article and its supplementary information files and from the corresponding author upon reasonable request.

## References

1. Arnold M, Rutherford MJ, Bardot A, et al. Progress in cancer survival, mortality, and incidence in seven high-income countries 1995–2014 (ICBP SURVMARK-2): a population-based study. The Lancet Oncology. 2019;20(11):1493–1505. doi:10.1016/S1470-2045(19)30456-5

2. Hudock NL, Mani K, Khunsriraksakul C, et al. Future trends in incidence and long-term survival of metastatic cancer in the United States. Commun Med (Lond). 2023;3:76. doi:10.1038/s43856-023-00304-x

3. Copur M, Suresh Manapuram MD. Multiple Primary Tumors Over a Lifetime. 2019;33. Accessed September 6, 2023. https://www.cancernetwork.com/view/multiple-primary-tumors-over-lifetime

4. Soerjomataram I, Coebergh JW. Epidemiology of Multiple Primary Cancers. In: Verma M, ed. Cancer Epidemiology. Methods in Molecular Biology. Humana Press; 2009:85–105. doi:10.1007/978-1-59745-416-2_5

5. Ye Y, Neil AL, Wills KE, Venn AJ. Temporal trends in the risk of developing multiple primary cancers: a systematic review. BMC Cancer. 2016;16(1):849. doi:10.1186/s12885-016-2876-y

6. Vogt A, Schmid S, Heinimann K, et al. Multiple primary tumours: challenges and approaches, a review. ESMO Open. 2017;2(2):e000172. doi:10.1136/esmoopen-2017-000172

7. Bychkovsky BL, Lo MT, Yussuf A, et al. Prevalence and spectrum of pathogenic variants among patients with multiple primary cancers evaluated by clinical characteristics. Cancer. 2022;128(6):1275–1283. doi:10.1002/cncr.34056

8. Cavazos TB, Kachuri L, Graff RE, et al. Assessment of genetic susceptibility to multiple primary cancers through whole-exome sequencing in two large multi-ancestry studies. BMC Med. 2022;20(1):332. doi:10.1186/s12916-022-02535-6

9. Chan GHJ, Ong PY, Low JJH, et al. Clinical genetic testing outcome with multi-gene panel in Asian patients with multiple primary cancers. Oncotarget. 2018;9(55):30649–30660. doi:10.18632/oncotarget.25769

10. Whitworth J, Smith PS, Martin JE, et al. Comprehensive Cancer-Predisposition Gene Testing in an Adult Multiple Primary Tumor Series Shows a Broad Range of Deleterious Variants and Atypical Tumor Phenotypes. Am J Hum Genet. 2018;103(1):3–18. doi:10.1016/j.ajhg.2018.04.013

11. Pritchard AL, Johansson PA, Nathan V, et al. Germline mutations in candidate predisposition genes in individuals with cutaneous melanoma and at least two independent additional primary cancers. Haass NK, ed. PLoS ONE. 2018;13(4):e0194098. doi:10.1371/journal.pone.0194098

12. Rashkin SR, Graff RE, Kachuri L, et al. Pan-cancer study detects genetic risk variants and shared genetic basis in two large cohorts. Nat Commun. 2020;11:4423. doi:10.1038/s41467-020-18246-6

13. Wu YH, Graff RE, Passarelli MN, et al. Identification of pleiotropic cancer susceptibility variants from genome-wide association studies reveals functional characteristics. Cancer Epidemiol Biomarkers Prev. 2018;27(1):75–85. doi:10.1158/1055-9965.EPI-17-0516

14. Lindström S, Wang L, Feng H, et al. Genome-wide analyses characterize shared heritability among cancers and identify novel cancer susceptibility regions. J Natl Cancer Inst. 2023;115(6):712–732. doi:10.1093/jnci/djad043

15. Guo H, Cao W, Zhu Y, Li T, Hu B. A genome-wide cross-cancer meta-analysis highlights the shared genetic links of five solid cancers. Frontiers in Microbiology. 2023;14. Accessed July 5, 2023. https://www.frontiersin.org/articles/10.3389/fmicb.2023.1116592

16. Fehringer G, Kraft P, Pharoah PD, et al. Cross-Cancer Genome-Wide Analysis of Lung, Ovary, Breast, Prostate, and Colorectal Cancer Reveals Novel Pleiotropic Associations. Cancer Res. 2016;76(17):5103–5114. doi:10.1158/0008-5472.CAN-15-2980

17. Jiang X, Finucane HK, Schumacher FR, et al. Shared heritability and functional enrichment across six solid cancers. Nat Commun. 2019;10(1):431. doi:10.1038/s41467-018-08054-4

18. Bycroft C, Freeman C, Petkova D, et al. The UK Biobank resource with deep phenotyping and genomic data. Nature. 2018;562(7726):203–209. doi:10.1038/s41586-018-0579-z

19. Hoffmann TJ, Van Den Eeden SK, Sakoda LC, et al. A large multiethnic genome-wide association study of prostate cancer identifies novel risk variants and substantial ethnic differences. Cancer Discov. 2015;5(8):878–891. doi:10.1158/2159-8290.CD-15-0315

20. Graff RE, Cavazos TB, Thai KK, et al. Cross-cancer evaluation of polygenic risk scores for 16 cancer types in two large cohorts. Nat Commun. 2021;12(1):970. doi:10.1038/s41467-021-21288-z

21. Adams S, Bs MB, Cheney H, et al. SEER Program Coding and Staging Manual 2021. Published online 2021.

22. Harris NL, Jaffe ES, Diebold J, et al. The World Health Organization classification of neoplastic diseases of the hematopoietic and lymphoid tissues. Report of the Clinical Advisory Committee meeting, Airlie House, Virginia, November, 1997. Ann Oncol. 1999;10(12):1419–1432. doi:10.1023/a:1008375931236

23. Manichaikul et al. A. Robust relationship inference in genome-wide association studies | Bioinformatics | Oxford Academic. Accessed February 19, 2024. https://academic.oup.com/bioinformatics/article/26/22/2867/228512

24. Hoffmann TJ, Kvale MN, Hesselson SE, et al. Next generation genome-wide association tool: Design and coverage of a high-throughput European-optimized SNP array. Genomics. 2011;98(2):79–89. doi:10.1016/j.ygeno.2011.04.005

25. Hoffmann TJ, Zhan Y, Kvale MN, et al. Design and coverage of high throughput genotyping arrays optimized for individuals of East Asian, African American, and Latino race/ethnicity using imputation and a novel hybrid SNP selection algorithm. Genomics. 2011;98(6):422–430. doi:10.1016/j.ygeno.2011.08.007

26. Kvale MN, Hesselson S, Hoffmann TJ, et al. Genotyping Informatics and Quality Control for 100,000 Subjects in the Genetic Epidemiology Research on Adult Health and Aging (GERA) Cohort. Genetics. 2015;200(4):1051–1060. doi:10.1534/genetics.115.178905

27. Delaneau O, Marchini J, Zagury JF. A linear complexity phasing method for thousands of genomes. Nat Methods. 2011;9(2):179–181. doi:10.1038/nmeth.1785

28. Howie BN, Donnelly P, Marchini J. A Flexible and Accurate Genotype Imputation Method for the Next Generation of Genome-Wide Association Studies. Schork NJ, ed. PLoS Genet. 2009;5(6):e1000529. doi:10.1371/journal.pgen.1000529

29. Banda Y, Kvale MN, Hoffmann TJ, et al. Characterizing Race/Ethnicity and Genetic Ancestry for 100,000 Subjects in the Genetic Epidemiology Research on Adult Health and Aging (GERA) Cohort. Genetics. 2015;200(4):1285–1295. doi:10.1534/genetics.115.178616

30. Price AL, Patterson NJ, Plenge RM, Weinblatt ME, Shadick NA, Reich D. Principal components analysis corrects for stratification in genome-wide association studies. Nat Genet. 2006;38(8):904–909. doi:10.1038/ng1847

31. Willer CJ, Li Y, Abecasis GR. METAL: fast and efficient meta-analysis of genomewide association scans. Bioinformatics. 2010;26(17):2190–2191. doi:10.1093/bioinformatics/btq340

32. Barbeira AN, Dickinson SP, Bonazzola R, et al. Exploring the phenotypic consequences of tissue specific gene expression variation inferred from GWAS summary statistics. Nat Commun. 2018;9(1):1825. doi:10.1038/s41467-018-03621-1

33. Barbeira AN, Pividori M, Zheng J, Wheeler HE, Nicolae DL, Im HK. Integrating predicted transcriptome from multiple tissues improves association detection. PLOS Genetics. 2019;15(1):e1007889. doi:10.1371/journal.pgen.1007889

34. Barbeira AN, Melia OJ, Liang Y, et al. Fine-mapping and QTL tissue-sharing information improves the reliability of causal gene identification. Genet Epidemiol. 2020;44(8):854–867. doi:10.1002/gepi.22346

35. Dorajoo R, Chang X, Gurung RL, et al. Loci for human leukocyte telomere length in the Singaporean Chinese population and trans-ethnic genetic studies. Nat Commun. 2019;10(1):2491. doi:10.1038/s41467-019-10443-2

36. Gudmundsson J, Thorleifsson G, Sigurdsson JK, et al. A genome-wide association study yields five novel thyroid cancer risk loci. Nat Commun. 2017;8:14517. doi:10.1038/ncomms14517

37. Landi MT, Bishop DT, MacGregor S, et al. Genome-wide association meta-analyses combining multiple risk phenotypes provide insights into the genetic architecture of cutaneous melanoma susceptibility. Nat Genet. 2020;52(5):494–504. doi:10.1038/s41588-020-0611-8

38. McKay JD, Hung RJ, Han Y, et al. Large-scale association analysis identifies new lung cancer susceptibility loci and heterogeneity in genetic susceptibility across histological subtypes. Nat Genet. 2017;49(7):1126–1132. doi:10.1038/ng.3892

39. Phelan CM, Kuchenbaecker KB, Tyrer JP, et al. Identification of 12 new susceptibility loci for different histotypes of epithelial ovarian cancer. Nat Genet. 2017;49(5):680–691. doi:10.1038/ng.3826

40. Scelo G, Purdue MP, Brown KM, et al. Genome-wide association study identifies multiple risk loci for renal cell carcinoma. Nat Commun. 2017;8(1):15724. doi:10.1038/ncomms15724

41. Son HY, Hwangbo Y, Yoo SK, et al. Genome-wide association and expression quantitative trait loci studies identify multiple susceptibility loci for thyroid cancer. Nat Commun. 2017;8(1):15966. doi:10.1038/ncomms15966

42. Whiffin N, Hosking FJ, Farrington SM, et al. Identification of susceptibility loci for colorectal cancer in a genome-wide meta-analysis. Human Molecular Genetics. 2014;23(17):4729–4737. doi:10.1093/hmg/ddu177

43. Peña-Chilet M, Blanquer-Maceiras M, Ibarrola-Villava M, et al. Genetic variants in PARP1 (rs3219090) and IRF4(rs12203592) genes associated with melanoma susceptibility in a Spanish population. BMC Cancer. 2013;13(1):160. doi:10.1186/1471-2407-13-160

44. Do TN, Ucisik-Akkaya E, Davis CF, Morrison BA, Dorak MT. An intronic polymorphism of IRF4 gene influences gene transcription in vitro and shows a risk association with childhood acute lymphoblastic leukemia in males. Biochim Biophys Acta. 2010;1802(2):292–300. doi:10.1016/j.bbadis.2009.10.015

45. Qian Y, Du Z, Xing Y, Zhou T, Chen T, Shi M. Interferon regulatory factor 4 (IRF4) is overexpressed in human nonlZIsmall cell lung cancer (NSCLC) and activates the Notch signaling pathway. Mol Med Rep. 2017;16(5):6034–6040. doi:10.3892/mmr.2017.7319

46. Rauch DA, Olson SL, Harding JC, et al. Interferon regulatory factor 4 as a therapeutic target in adult T-cell leukemia lymphoma. Retrovirology. 2020;17(1):27. doi:10.1186/s12977-020-00535-z

47. Byun J, Han Y, Li Y, et al. Cross-ancestry genome-wide meta-analysis of 61,047 cases and 947,237 controls identifies new susceptibility loci contributing to lung cancer. Nat Genet. 2022;54(8):1167–1177. doi:10.1038/s41588-022-01115-x

48. Levy D, Neuhausen SL, Hunt SC, et al. Genome-wide association identifies OBFC1 as a locus involved in human leukocyte telomere biology. Proceedings of the National Academy of Sciences. 2010;107(20):9293–9298. doi:10.1073/pnas.0911494107

49. Taub MA, Conomos MP, Keener R, et al. Genetic determinants of telomere length from 109,122 ancestrally diverse whole-genome sequences in TOPMed. Cell Genomics. 2022;2(1):100084. doi:10.1016/j.xgen.2021.100084

50. Law PJ, Sud A, Mitchell JS, et al. Genome-wide association analysis of chronic lymphocytic leukaemia, Hodgkin lymphoma and multiple myeloma identifies pleiotropic risk loci. Sci Rep. 2017;7(1):41071. doi:10.1038/srep41071

51. Codd V, Mangino M, van der Harst P, et al. Common variants near TERC are associated with mean telomere length. Nat Genet. 2010;42(3):197–199. doi:10.1038/ng.532

52. Soerensen M, Thinggaard M, Nygaard M, et al. Genetic variation in TERT and TERC and human leukocyte telomere length and longevity: a cross-sectional and longitudinal analysis. Aging Cell. 2012;11(2):223–227. doi:10.1111/j.1474-9726.2011.00775.x

53. Jones AM, Beggs AD, Carvajal-Carmona L, et al. TERC polymorphisms are associated both with susceptibility to colorectal cancer and with longer telomeres. Gut. 2012;61(2):248–254. doi:10.1136/gut.2011.239772

54. Walsh KM, Codd V, Smirnov IV, et al. Variants near TERT and TERC influencing telomere length are associated with high-grade glioma risk. Nat Genet. 2014;46(7):731–735. doi:10.1038/ng.3004

55. Simpson BS, Camacho N, Luxton HJ, et al. Genetic alterations in the 3q26.31-32 locus confer an aggressive prostate cancer phenotype. Commun Biol. 2020;3(1):1–10. doi:10.1038/s42003-020-01175-x

56. Houlston RS, Cheadle J, Dobbins SE, et al. Meta-analysis of three genome-wide association studies identifies susceptibility loci for colorectal cancer at 1q41, 3q26.2, 12q13.13 and 20q13.33. Nat Genet. 2010;42(11):973–977. doi:10.1038/ng.670

57. Figueroa JD, Ye Y, Siddiq A, et al. Genome-wide association study identifies multiple loci associated with bladder cancer risk. Human Molecular Genetics. 2014;23(5):1387–1398. doi:10.1093/hmg/ddt519

58. Polat F, Yilmaz M, Budak Diler S. The Association of MYNN and TERC Gene Polymorphisms and Bladder Cancer in a Turkish Population. Urol J. 2019;16(1):50–55. doi:10.22037/uj.v0i0.4083

59. Wang M, Chu H, Lv Q, et al. Cumulative effect of genome-wide association study-identified genetic variants for bladder cancer. International Journal of Cancer. 2014;135(11):2653–2660. doi:10.1002/ijc.28898

60. Campa D, Martino A, Varkonyi J, et al. Risk of multiple myeloma is associated with polymorphisms within telomerase genes and telomere length. Int J Cancer. 2015;136(5):E351–358. doi:10.1002/ijc.29101

61. Giaccherini M, Macauda A, Orciuolo E, et al. Genetically determined telomere length and multiple myeloma risk and outcome. Blood Cancer J. 2021;11(4):1–10. doi:10.1038/s41408-021-00462-y

62. Qin N, Li Y, Wang C, et al. Comprehensive functional annotation of susceptibility variants identifies genetic heterogeneity between lung adenocarcinoma and squamous cell carcinoma. Front Med. 2021;15(2):275–291. doi:10.1007/s11684-020-0779-4

63. Comiskey DF, He H, Liyanarachchi S, et al. Variants in LRRC34 reveal distinct mechanisms for predisposition to papillary thyroid carcinoma. J Med Genet. 2020;57(8):519–527. doi:10.1136/jmedgenet-2019-106554

64. Zeid D, Mooney-Leber S, Seemiller LR, Goldberg LR, Gould TJ. Terc Gene Cluster Variants Predict Liver Telomere Length in Mice. Cells. 2021;10(10):2623. doi:10.3390/cells10102623

65. Li C, Stoma S, Lotta LA, et al. Genome-wide Association Analysis in Humans Links Nucleotide Metabolism to Leukocyte Telomere Length. Am J Hum Genet. 2020;106(3):389–404. doi:10.1016/j.ajhg.2020.02.006

66. Akincilar SC, Chan CHT, Ng QF, Fidan K, Tergaonkar V. Non-canonical roles of canonical telomere binding proteins in cancers. Cell Mol Life Sci. 2021;78(9):4235–4257. doi:10.1007/s00018-021-03783-0

67. Teng Y, Huang DQ, Li RX, Yi C, Zhan YQ. Association Between Telomere Length and Risk of Lung Cancer in an Asian Population: A Mendelian Randomization Study. World J Oncol. 2023;14(4):277–284. doi:10.14740/wjon1624

68. Telomeres Mendelian Randomization Collaboration, Haycock PC, Burgess S, et al. Association Between Telomere Length and Risk of Cancer and Non-Neoplastic Diseases: A Mendelian Randomization Study. JAMA Oncol. 2017;3(5):636–651. doi:10.1001/jamaoncol.2016.5945

69. Kachuri L, Saarela O, Bojesen SE, et al. Mendelian Randomization and mediation analysis of leukocyte telomere length and risk of lung and head and neck cancers. Int J Epidemiol. 2019;48(3):751–766. doi:10.1093/ije/dyy140

70. McNally EJ, Luncsford PJ, Armanios M. Long telomeres and cancer risk: the price of cellular immortality. J Clin Invest. 2019;129(9):3474–3481. doi:10.1172/JCI120851

71. Saunders CN, Kinnersley B, Culliford R, Cornish AJ, Law PJ, Houlston RS. Relationship between genetically determined telomere length and glioma risk. Neuro Oncol. 2022;24(2):171–181. doi:10.1093/neuonc/noab208

72. Demanelis K, Jasmine F, Chen LS, et al. Determinants of telomere length across human tissues. Science. 2020;369(6509):eaaz6876. doi:10.1126/science.aaz6876

73. Stewart JA, Wang Y, Ackerson SM, Schuck PL. Emerging roles of CST in maintaining genome stability and human disease. Front Biosci (Landmark Ed). 2018;23:1564–1586.

74. Artandi SE, DePinho RA. Telomeres and telomerase in cancer. Carcinogenesis. 2010;31(1):9–18. doi:10.1093/carcin/bgp268

75. Di Giovannantonio M, Harris BH, Zhang P, et al. Heritable genetic variants in key cancer genes link cancer risk with anthropometric traits. J Med Genet. 2021;58(6):392–399. doi:10.1136/jmedgenet-2019-106799

76. Eiholzer RA, Mehta S, Kazantseva M, et al. Intronic TP53 Polymorphisms Are Associated with Increased Δ133TP53 Transcript, Immune Infiltration and Cancer Risk. Cancers (Basel). 2020;12(9):2472. doi:10.3390/cancers12092472

77. Steffens Reinhardt L, Groen K, Xavier A, Avery-Kiejda KA. p53 Dysregulation in Breast Cancer: Insights on Mutations in the TP53 Network and p53 Isoform Expression. Int J Mol Sci. 2023;24(12):10078. doi:10.3390/ijms241210078

78. Diskin SJ, Capasso M, Diamond M, et al. Rare variants in TP53 and susceptibility to neuroblastoma. J Natl Cancer Inst. 2014;106(4):dju047. doi:10.1093/jnci/dju047

79. Han J, Qureshi AA, Nan H, et al. A Germline Variant in the Interferon Regulatory Factor 4 Gene as a Novel Skin Cancer Risk Locus. Cancer Res. 2011;71(5):1533–1539. doi:10.1158/0008-5472.CAN-10-1818

80. Wang S, Yan Q, Chen P, Zhao P, Gu A. Association of interferon regulatory factor 4 gene polymorphisms rs12203592 and rs872071 with skin cancer and haematological malignancies susceptibility: a meta-analysis of 19 case–control studies. BMC Cancer. 2014;14(1):410. doi:10.1186/1471-2407-14-410

81. Wang J, Clay-Gilmour AI, Karaesmen E, et al. Genome-Wide Association Analyses Identify Variants in IRF4 Associated With Acute Myeloid Leukemia and Myelodysplastic Syndrome Susceptibility. Frontiers in Genetics. 2021;12. Accessed February 17, 2024. https://www.frontiersin.org/journals/genetics/articles/10.3389/fgene.2021.554948

82. Wong RWJ, Ong JZL, Theardy MS, Sanda T. IRF4 as an Oncogenic Master Transcription Factor. Cancers (Basel). 2022;14(17):4314. doi:10.3390/cancers14174314

83. Lu J, Liang T, Li P, Yin Q. Regulatory effects of IRF4 on immune cells in the tumor microenvironment. Frontiers in Immunology. 2023;14. Accessed February 17, 2024. https://www.frontiersin.org/journals/immunology/articles/10.3389/fimmu.2023.1086803

84. Shaffer AL, Emre NCT, Romesser PB, Staudt LM. IRF4: Immunity. Malignancy! Therapy? Clinical Cancer Research. 2009;15(9):2954–2961. doi:10.1158/1078-0432.CCR-08-1845

85. Li X, Zhai S, Zhang J, et al. Interferon Regulatory Factor 4 Correlated With Immune Cells Infiltration Could Predict Prognosis for Patients With Lung Adenocarcinoma. Front Oncol. 2021;11:698465. doi:10.3389/fonc.2021.698465

86. Heimes AS, Madjar K, Edlund K, et al. Prognostic significance of interferon regulating factor 4 (IRF4) in node-negative breast cancer. J Cancer Res Clin Oncol. 2017;143(7):1123–1131. doi:10.1007/s00432-017-2377-7

87. Lagou S, Grapsa D, Syrigos N, Bamias G. The Role of Decoy Receptor DcR3 in Gastrointestinal Malignancy. Cancer Diagn Progn. 2022;2(4):411–421. doi:10.21873/cdp.10124

88. Liang Q lian, Wang B rong, Li G hong. DcR3 and survivin are highly expressed in colorectal carcinoma and closely correlated to its clinicopathologic parameters. J Zhejiang Univ Sci B. 2009;10(9):675–682. doi:10.1631/jzus.B0920077

89. Yu W, Xu YC, Tao Y, et al. DcR3 regulates the growth and metastatic potential of SW480 colon cancer cells. Oncol Rep. 2013;30(6):2741–2748. doi:10.3892/or.2013.2769

90. Zhong M, Qiu X, Liu Y, et al. TIPE Regulates DcR3 Expression and Function by Activating the PI3K/AKT Signaling Pathway in CRC. Frontiers in Oncology. 2021;10. Accessed February 20, 2024. https://www.frontiersin.org/journals/oncology/articles/10.3389/fonc.2020.623048

91. Chen J, Guo XZ, Li HY, Zhao JJ, Xu WD. Dendritic cells engineered to secrete anti-DcR3 antibody augment cytotoxic T lymphocyte response against pancreatic cancer in vitro. World J Gastroenterol. 2017;23(5):817–829. doi:10.3748/wjg.v23.i5.817

92. Wang W, Zhang M, Sun W, et al. Reduction of Decoy Receptor 3 Enhances TRAIL-Mediated Apoptosis in Pancreatic Cancer. PLOS ONE. 2013;8(10):e74272. doi:10.1371/journal.pone.0074272

93. Wei Y, Chen X, Yang J, et al. DcR3 promotes proliferation and invasion of pancreatic cancer via a DcR3/STAT1/IRF1 feedback loop. Am J Cancer Res. 2019;9(12):2618–2633.

94. Zhou J, Song S, Li D, et al. Decoy receptor 3 (DcR3) overexpression predicts the prognosis and pN2 in pancreatic head carcinoma. World Journal of Surgical Oncology. 2014;12(1):52. doi:10.1186/1477-7819-12-52

95. Connor JP, Felder M. Ascites from epithelial ovarian cancer contain high levels of functional decoy receptor 3 (DcR3) and is associated with platinum resistance. Gynecologic Oncology. 2008;111(2):330–335. doi:10.1016/j.ygyno.2008.07.012

96. Jiang M, Lin X, He R, et al. Decoy Receptor 3 (DcR3) as a Biomarker of Tumor Deterioration in Female Reproductive Cancers: A Meta-Analysis. Med Sci Monit. 2016;22:1850–1857. doi:10.12659/MSM.896226

97. Chang YH, Wang PH, Chen YJ. Expression of DcR3 in ovarian cancer and clinicopathological implication. Gynecologic Oncology. 2020;159:341–342. doi:10.1016/j.ygyno.2020.05.631

98. Liang C, Xu Y, Li G, et al. Downregulation of DcR3 sensitizes hepatocellular carcinoma cells to TRAIL-induced apoptosis. Onco Targets Ther. 2017;10:417–428. doi:10.2147/OTT.S127202

99. Chen G, Luo D. Expression of decoy receptor 3 in liver tissue microarrays. Natl Med J India. 2008;21(6):275–278.

100. Hsieh SL, Lin WW. Decoy receptor 3: an endogenous immunomodulator in cancer growth and inflammatory reactions. Journal of Biomedical Science. 2017;24(1):39. doi:10.1186/s12929-017-0347-7

101. You RI, Chang YC, Chen PM, et al. Apoptosis of dendritic cells induced by decoy receptor 3 (DcR3). Blood. 2008;111(3):1480–1488. doi:10.1182/blood-2007-09-114850

102. Yang CR, Hsieh SL, Teng CM, Ho FM, Su WL, Lin WW. Soluble decoy receptor 3 induces angiogenesis by neutralization of TL1A, a cytokine belonging to tumor necrosis factor superfamily and exhibiting angiostatic action. Cancer Res. 2004;64(3):1122–1129. doi:10.1158/0008-5472.can-03-0609

103. Rashid M ur, Lorzadeh S, Gao A, Ghavami S, Coombs KM. PSMA2 knockdown impacts expression of proteins involved in immune and cellular stress responses in human lung cells. Biochimica et Biophysica Acta (BBA) - Molecular Basis of Disease. 2023;1869(2):166617. doi:10.1016/j.bbadis.2022.166617

104. Tanaka K, Tsurumi C. The 26S proteasome: subunits and functions. Mol Biol Rep. 1997;24(1):3–11. doi:10.1023/A:1006876904158

105. Chiao CC, Liu YH, Phan NN, et al. Prognostic and Genomic Analysis of Proteasome 20S Subunit Alpha (PSMA) Family Members in Breast Cancer. Diagnostics (Basel). 2021;11(12):2220. doi:10.3390/diagnostics11122220

106. Qi J, Hu Z, Liu S, et al. Comprehensively Analyzed Macrophage-Regulated Genes Indicate That PSMA2 Promotes Colorectal Cancer Progression. Front Oncol. 2021;10. doi:10.3389/fonc.2020.618902

107. Zhang Y, Xiang Z, Chen L, Deng X, Liu H, Peng X. PSMA2 promotes glioma proliferation and migration via EMT. Pathol Res Pract. 2024;256:155278. doi:10.1016/j.prp.2024.155278

