## Supplementary Figures for "Unraveling the genetic landscape of susceptibility to multiple primary cancers"

Supplementary Figure 1: Circos plot showing the pairs of cancer diagnoses with at least 50 individuals in the (A) UK Biobank study and (B) GERA study. Each connection reflects the number of cases with both linked primary cancers, where the color of the line shows the first cancer site diagnosed.

(A) UK Biobank

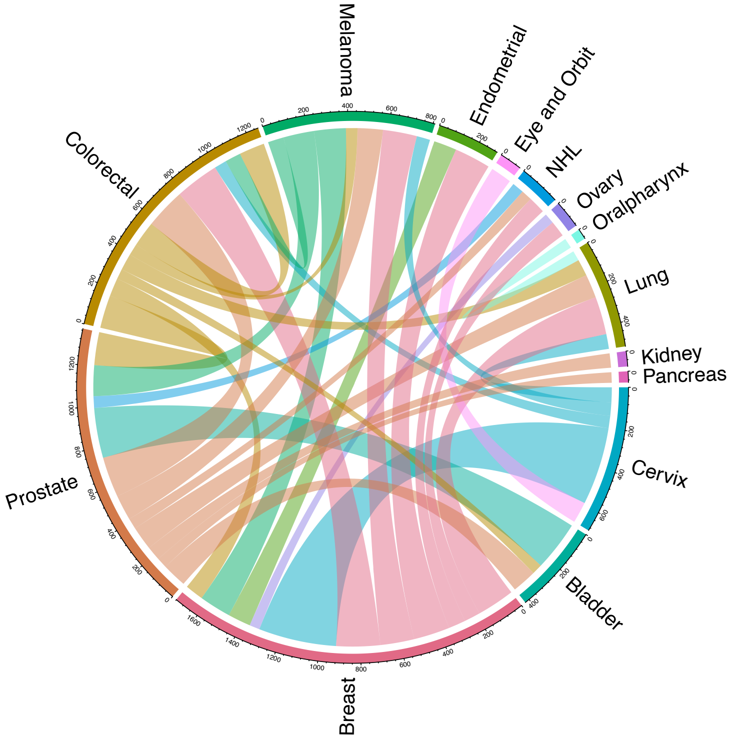


(B) GERA study

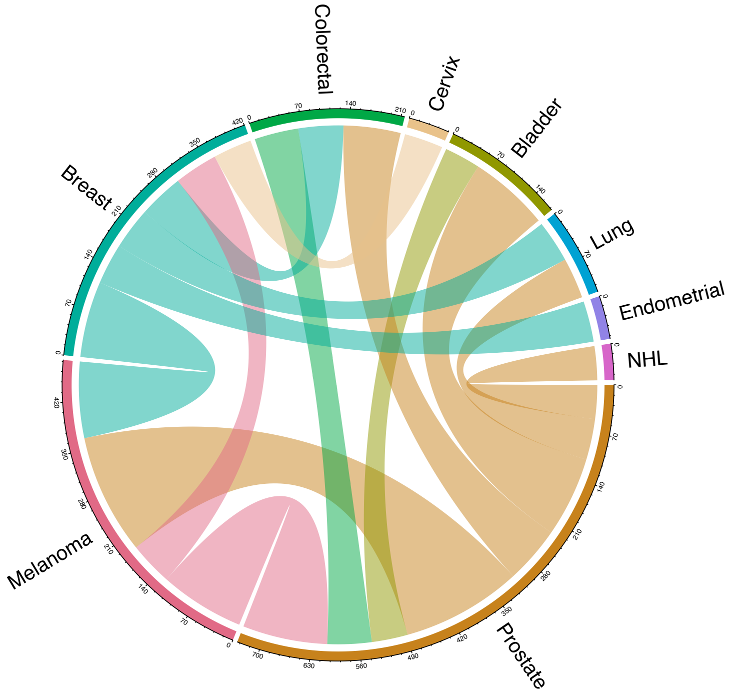

Supplementary Figure 2: Manhattan plot highlighting the lead chromosomal regions ($P<5\times10^{-8}$) from the genome-wide association study of multiple invasive cancers (n=8,673) versus cancer-free controls (n=420,944). The solid black line signifies the genome-wide significance threshold ($P<5\times10^{-8}$), whereas the dashed black represents a suggestive significance threshold of $P<1\times10^{-6}$.

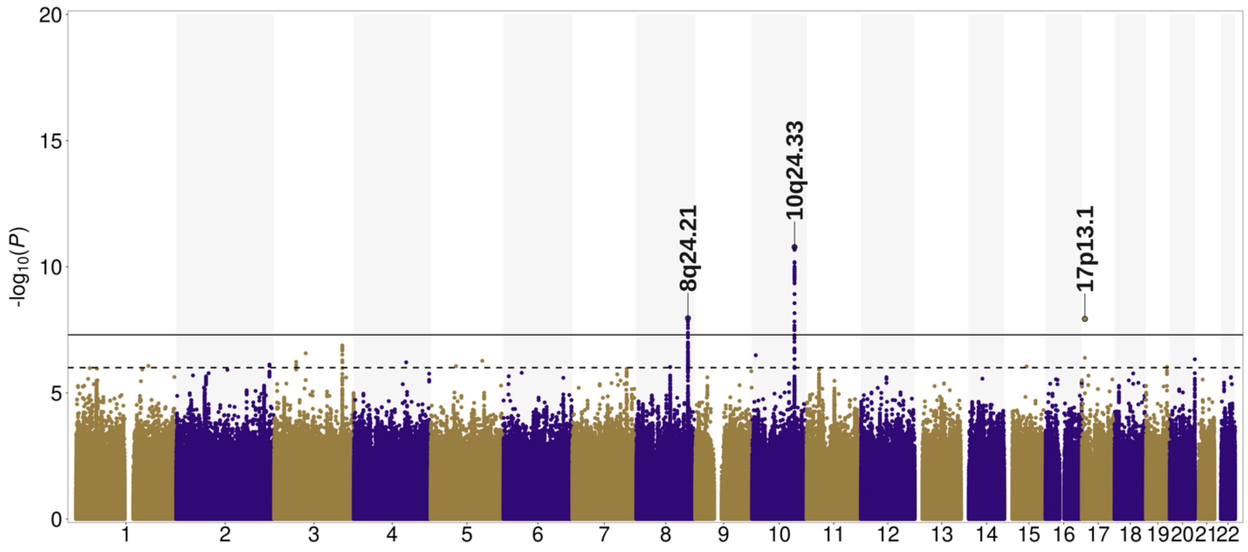

Supplementary Figure 3: Multi-tissue transcriptome-wide association study (TWAS) results for (A) multiple cancers versus single cancer and (B) multiple invasive cancers versus single invasive cancer. The top panel of the Miami plots depicts the associations for genes with mean Z scores > 0, and the bottom panel shows genes with mean Z scores ≤ 0. The threshold for statistical significance was determined based on the Bonferroni correction for 22,244 genes tested ($P<2.3\times10^{-6}$, dashed black line), while the suggestive significance threshold was set at $P<1\times10^{-4}$ (dashed blue line).

(A) TWAS of multiple cancers
(10,983 cases and 84,475 single cancer cases as controls)

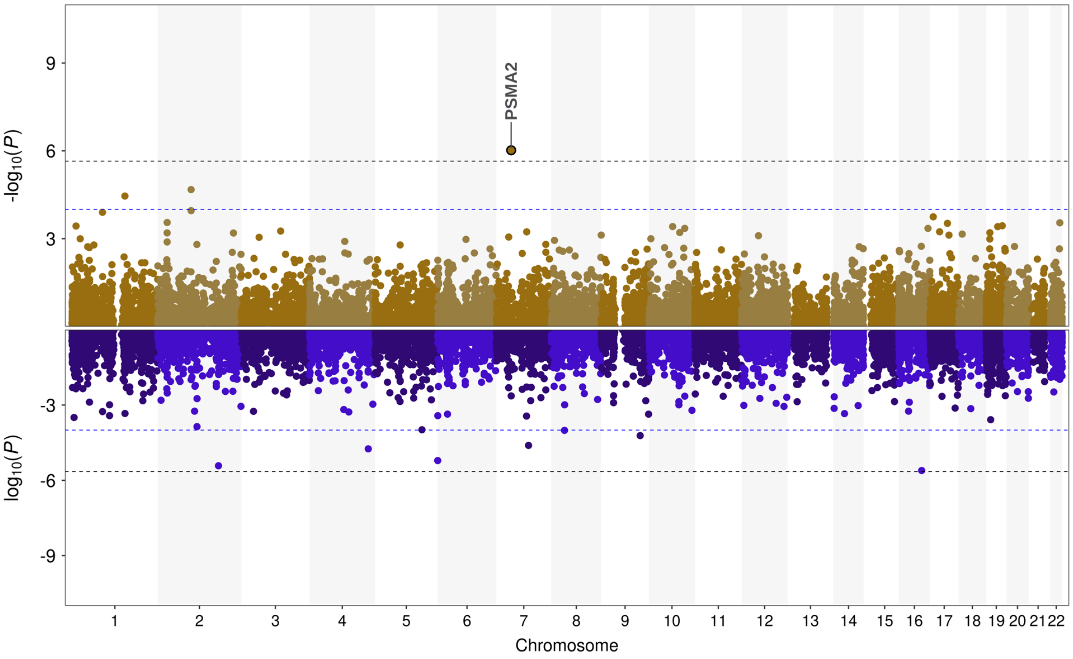


(B) TWAS of multiple invasive cancers
(8,673 cases and 72,686 single invasive cancer cases as controls)

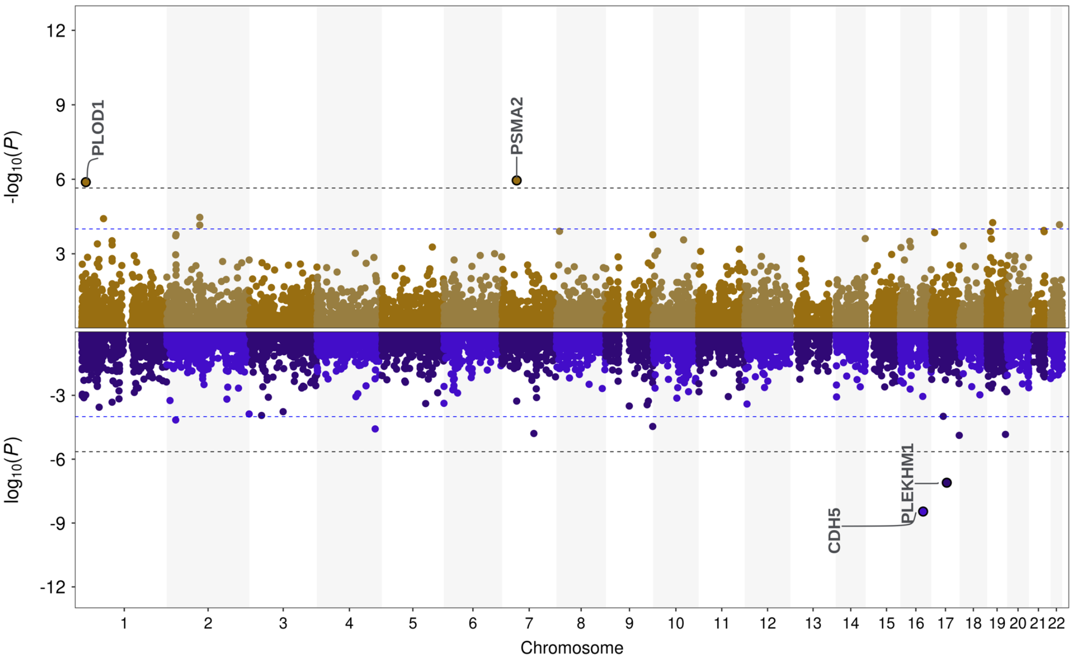
